# Rurality Modifies the Association Between Symptoms and the Diagnosis of Amyotrophic Lateral Sclerosis

**DOI:** 10.1101/2023.10.13.23297024

**Authors:** Alexander A Hart, Andrea Swenson, Nandakumar S. Narayanan, Jacob E. Simmering

**Affiliations:** Department of Neurology, University of Michigan, Ann Arbor, MI, 48109; Department of Neurology, University of Iowa, Iowa City, IA, 52242; Department of Internal Medicine, University of Iowa, Iowa City, IA, 52242

**Keywords:** Disparities, Neuromuscular, Quality Improvement

## Abstract

**Objective:** We utilized national claims-based data to identify the change in hazard of diagnosis of ALS following a possible ALS-related diagnosis (e.g., falls) and whether the change in hazard varies whether the patient lives in an urban or rural area.

**Methods:** Health insurance claims data from both the commercial insurance market and Medicare supplemental market were obtained from the Merative MarketScan Commercial Claims and Encounters and Medicare Coordination of Benefits databases. Individuals with a diagnosis of ALS were identified and matched on age, sex, and enrollment period to individuals without ALS. For all individuals, inciting events such as falls, muscle related symptoms, or bulbar symptoms were also extracted. We then used fixed-effects regression to estimate the risk of being diagnosed with ALS following one of these events controlling for urban-rural status. Additionally, we utilized interaction terms to evaluate the effect of rurality on odds of diagnosis.

**Results:** 19,126 individuals with ALS were included with 96,126 controls. Patients with ALS were more likely than matched controls to live in an urban area (87 vs 85%). Of those with ALS 84% had a symptom code preceding their diagnosis as compared to 51% in the general population. The association between having any symptoms and future diagnosis of ALS remained statistically significant after adjustment for confounders and an odds ratio of nearly 5. Odds ratios for the individual symptoms varied from 1.2 to over 10. In all models, living in an urban area was associated with increased odds of diagnosis with ALS while the effect of having a symptom was smaller among urban dwellers. Urban dwellers who are diagnosed with ALS are diagnosed at younger ages.

**Conclusions:** Early diagnosis of ALS is vital for connecting patients with research and treatment options. These results suggest symptoms appear in the administrative health record potentially years before the diagnosis of ALS. Additionally, rural patients are diagnosed at later ages with a greater dependence on major symptoms than urban patients. These results highlight potential improvements for surveillance and screening for ALS.

## Introduction

Improving the quality of life and survival among people with amyotrophic lateral sclerosis (ALS) depends critically on early diagnosis. Yet symptoms later attributed to ALS may appear up to a year before diagnosis.(1) Understanding the timing and magnitude of potentially missed ALS symptoms may provide insight into potential prodromal signs and diagnostic red flags. Despite ALS symptoms being present, an ALS diagnosis may be missed when the symptoms are overlooked or ambigious. The potential for a missed symptom may be especially high for patients with limited access or long referral times to specialist care, such as those living in rural areas. Neurologists tend to practice in larger, urban centers (2, 3, 4) resulting in greater travel distances for rural patients.(5) These access-related barriers to care may cause rural patients to be more likely to have their symptoms missed.

In this study, we quantify the association between having ALS-like symptoms and later having a diagnosis of ALS while stratifying by whether a person lives in a metropolitan or non-metropolitan county. We hypothesized that the risk of ALS will be increased in people following ALS-like symptoms and further than the increase in risk will vary according to whether the patient lives in an metropolitan or non-metropolitan county.

## Materials and Methods

### Data Source

Data from the Merative MarketScan Commercial Claims and Encounters and the Medicare Coordination of Benefits databases from 2001 to 2021 were used for this study. These datasets include claims-based data from inpatient encounters, outpatient visits, and pharmacy claims.(6) Analysis of secondary deidentified data is not considered human subjects research.

### Case Definition

Using the healthcare insurance claims, we identified patients with ALS using ICD-9-CM (335.20) and ICD-10-CM (G12.21) diagnosis codes on any claim. Patients were also included if they had taken the ALS-specific medications edaravone or riluzole. Sodium phenylbutyrate-taurursodiol was not included as release occurred in September 2022. Medication dispensing events was performed by matching on National Drug Code numbers found using the 2015 Redbook. The date of diagnosis was defined as either the first claim with an ALS diagnosis or dispensing of ALS medication, whichever occurred first. We required at least one year of enrollment prior to the first ALS date to ensure we are identifying new cases.

We identified healthy controls – people without ALS – that were the same sex, age, and enrolled during the same period as the case. For instance, a male enrollee with ALS born in 1933, who first appears in the data in 2003, and is diagnosed with ALS in 2007 would be matched to a male born in 1933 and enrolled from 2003 to 2007 but who is never diagnosed with ALS. Up to five healthy controls were identified for each case.

### Symptom Identification

ALS symptoms were based on codes previously reported in an analysis of Medicare data.(7) Additionally, in our criteria we added falls as a symptom. The considered symptoms are classified into two major categories – bulbar or motor symptoms. We retained subdivisions within each as described previously.(7) We crosswalked the ICD-9-CM codes to ICD-10-CM using the Center for Medicare and Medicaid Services General Equivalence Crosswalk. The codes used are described in **Table 1**. We included only claims and diagnoses from before the ALS diagnosis date.

**Table 1:**
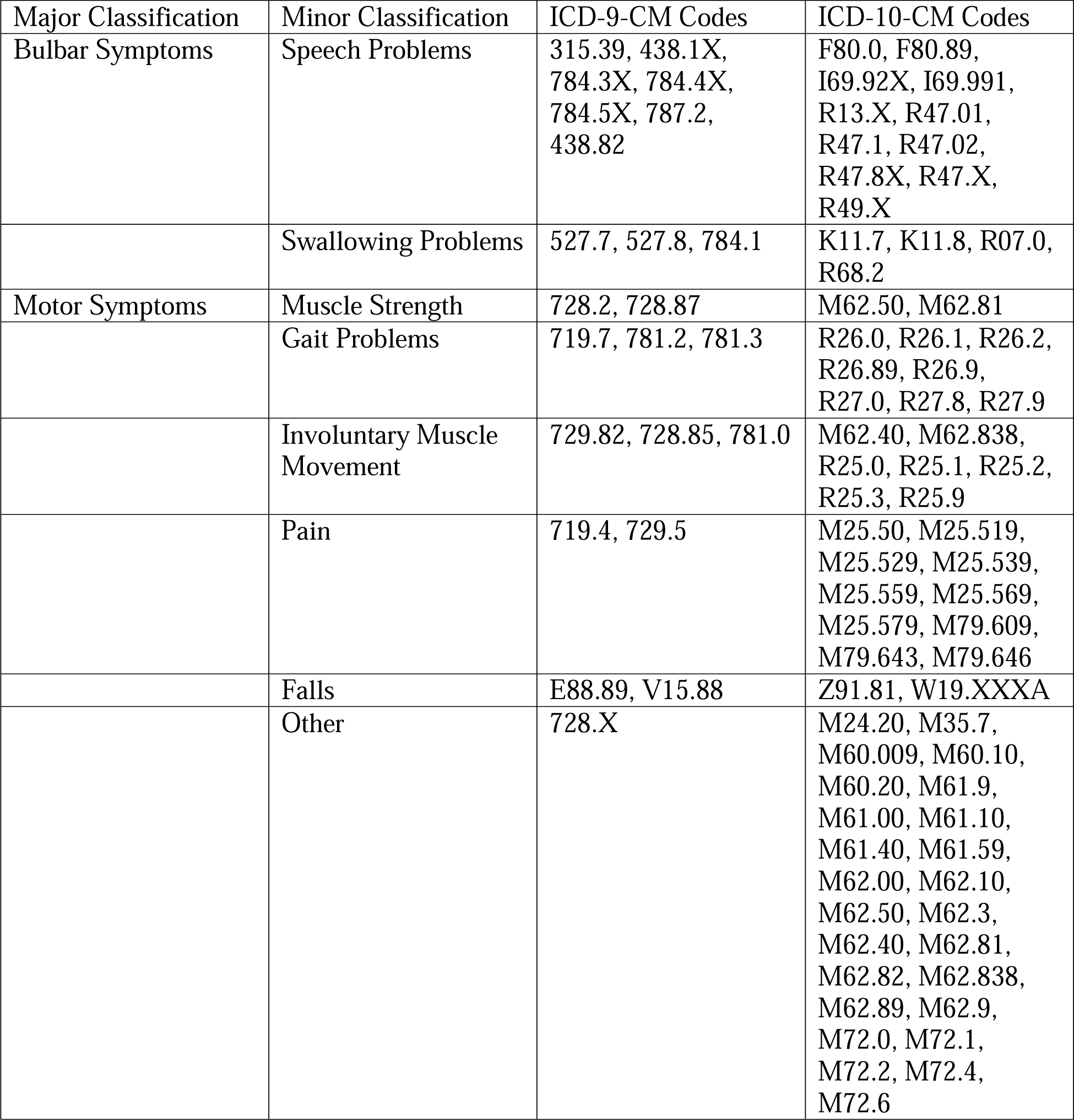
Diagnosis codes used to define ALS symptoms by origin and sub-classification.

### Urban/Rural Categorization

We defined all counties included in a metropolitan statistical area (MSA) as an urban area. MSAs are metro areas with populations of at least 50,000 and are defined by the Office of Management and Budget. Included in the MSA is the core city and closely linked surrounding area. If a person was ever reported as having lived in an MSA in Merative Marketscan, we coded them as metropolitan. Meanwhile, enrollees who never resided in an MSA were defined as rural. Roughly 85% of Americans and a similar percentage of enrollees in the Merative Marketscan database live in one of these MSAs.

### Primary Analysis

Our primary outcome was whether the person is ever diagnosed with or treated for ALS. Our primary exposures are 1) whether they had any possible ALS symptoms during the period before being diagnosed with ALS, 2) whether they resided in a metropolitan or non-metropolitan area, and 3) an interaction between these two exposures. We included adjustment for healthcare utilization during the pre-ALS period (rate of inpatient encounters, number of inpatient days, number of outpatient encounters), complexity (average number of diagnoses made per outpatient encounter), and comorbidities (the 30 AHRQ/Elixhauser comorbidity flags).(8, 9, 10) Estimation was performed using conditional logistic regression and included the sampling strata to account for matching on age, sex, and enrollment period.

### Secondary Analyses

We then conducted several secondary analyses to better understand the relationship between past ALS-like symptoms, urban dwelling, and the odds of ALS. First, we estimated “system” specific models to determine whether the association differed whether the symptoms were motor symptoms or bulbar origin. Second, we replaced the binary indicator for having any symptoms with the number of the eight defined symptoms that were observed to look for a dose-response relationship between ALS-like symptoms and the odds of getting an ALS diagnosis.

Third, since the different symptoms may have different relationships with the odds of developing ALS, we estimated eight symptom-specific models.

Additionally, we repeated our main analysis but eliminated cases where the symptom first occurred shortly before the diagnosis of ALS. A provider noting gait abnormalities and ordering a referral to neurology is an example of idealized diagnostic care – symptoms were noted and properly acted upon – but a delay necessarily exists between the symptoms being recognized and the definitive diagnosis. To address this possible concern, we implemented exclusion periods varying from 30 days to 2 years. A person with ALS-like symptoms within the exclusion period before they were diagnosed with ALS were removed from the analysis. For example, with a 180- day exclusion period, a person who first had an ALS-symptom within 180 days of getting diagnosed with ALS would be eliminated from the data. If an effect is found at a 180-day delay, for instance, it would suggest that the symptoms manifest in the administrative health record at least 6 months before the person is diagnosed with ALS.

Finally, given the lack of a previously established urban/rural difference in the incidence of ALS, we theorized any observed differences would likely result from different rates of detection and diagnosis due to greater access to care. If the true incidence of ALS in urban and rural areas is the same but people residing in urban areas are diagnosed earlier due to their greater access to care, we would expect ALS cases in urban areas to be diagnosed at younger ages than in rural areas. We performed a survival analysis to quantify differences in the age at diagnosis in people living in metropolitan or non-metropolitan counties and with or without symptoms. Specifically, we estimated the Kaplan-Meier survival function stratified by 1) living in a metropolitan or non-metropolitan country, 2) having or not having ALS symptoms, and 3) the interaction of these two variables. We quantified and tested these results using Cox proportional hazards regression.

All analysis were completed in R 4.2.2 (11) and the conditional logistic regression, Kaplan-Meier, and Cox proportional hazards models were estimated with the survival package.(12). For all regression models, standard errors were clustered by the sampling strata. In the Cox proportional hazards regression models, we used robust standard errors.

## Results

We identified 19,226 people newly diagnosed with ALS and 96,126 people without ALS matched based on age, sex, and enrollment period. People who will be diagnosed with ALS are slightly more likely to live in urban areas (87.0% vs 84.5%) and are much more likely to have one or more of the ALS-symptoms (83.7% vs 50.7%) than controls, **Table 2**. Differences were larger for higher number of symptoms (e.g., 25.5% of people who will develop ALS had 4 or more symptoms compared to only 4.7% of healthy controls) and specific symptoms (e.g., 35.1% of people who will develop ALS had a diagnosis of muscle weakness compared to 5.2% of healthy controls).

**Table 2:** Summary of Study Cohort. All values are reported as number (percent) when discrete or mean (inner quartile range) for continuous values. Age, sex, and enrollment time was including in the matching scheme and noted here (all with standardized mean differences below 0.001).

|  | Has ALS | Does Not Have ALS | Standardized Mean Difference |
| --- | --- | --- | --- |
| N | 19,226 | 96,126 |  |
| Age | 56 (47, 65) | 56 (47, 65) | Matched (< 0.001) |
| Female | 10,865 (56.5%) | 54,325 (56.5%) | Matched (< 0.001) |
| First Year in Data | 2007 (2003, 2010) | 2007 (2003, 2010) | Matched (< 0.001) |
| Last Year in Data | 2012 (2006, 2015) | 2012 (2008, 2015) | Matched (< 0.001) |
| Number of Outpatient Visits Per Year | 13.66 (7.26, 24.11) | 7.10 (2.89, 14.56) | 0.465 |
| Number of Inpatient Stays Per Year | 0.00 (0.00, 0.25) | 0.00 (0.00, 0.12) | 0.168 |
| Number of Diagnoses Per Outpatient Visit | 1.61 (1.29, 2.13) | 1.51 (1.21, 2.03) | 0.196 |
| Number of Inpatient Days Per Year | 0.00 (0.00, 2.50) | 0.00 (0.00, 1.50) | 0.127 |
| Elixhauser Comorbidities |  |  |  |
| HF | 1,771 (9.2%) | 8,899 (9.3%) | 0.002 |
| Valvular | 3,090 (16.1%) | 12,631 (13.1%) | 0.083 |
| Pulm. Hypertension | 755 (3.9%) | 3,107 (3.2%) | 0.037 |
| PVD | 3,035 (15.8%) | 12,089 (12.6%) | 0.092 |
| HTN | 10,604 (55.2%) | 48,410 (50.4%) | 0.096 |
| HTN with Complications | 2,338 (12.2%) | 11,065 (11.5%) | 0.020 |
| Paralysis | 2,219 (11.5%) | 2,065 (2.1%) | 0.379 |
| Neurological | 7,838 (40.8%) | 9,944 (10.3%) | 0.744 |
| COPD | 5,312 (27.6%) | 21,899 (22.8%) | 0.112 |
| DM | 5,312 (27.6%) | 20,001 (20.8%) | 0.005 |
| DM with Complications | 1,639 (8.5%) | 8,413 (8.8%) | 0.008 |
| Hypothyroidism | 3,550 (18.5%) | 13,543 (14.1%) | 0.119 |
| Renal Disease | 1,052 (5.5%) | 6,213 (6.5%) | 0.042 |
| Liver Disease | 1,079 (5.6%) | 4,626 (4.8%) | 0.036 |
| Peptic Ulcer Disorder | 127 (0.7%) | 491 (0.5%) | 0.020 |
| HIV | 51 (0.3%) | 188 (0.2%) | 0.015 |
| Lymphoma | 371 (1.9%) | 1,326 (1.4%) | 0.043 |
| Metastatic Cancer | 386 (2.0%) | 3,508 (3.6%) | 0.099 |
| Solid Tumor | 2,322 (12.1%) | 12,044 (12.5%) | 0.014 |
| Rheumatic Disease | 2,130 (11.1%) | 5,852 (6.1%) | 0.179 |
| Coagulopathy | 1,011 (5.3%) | 3,999 (4.2%) | 0.052 |
| Obesity | 2,218 (11.5%) | 11,063 (11.5%) | 0.001 |
| Weight Loss | 2,344 (12.2%) | 5,374 (5.6%) | 0.234 |
| Fluid and Electrolyte Disorder | 3,348 (17.4%) | 13,387 (13.9%) | 0.096 |
| Blood Loss | 397 (2.1%) | 2,005 (2.1%) | 0.001 |
| Anemia | 4,267 (22.2%) | 16,207 (16.9%) | 0.135 |
| Alcohol Abuse | 491 (2.6%) | 2,022 (2.1%) | 0.030 |
| Drug Abuse | 397 (2.1%) | 1,292 (1.3%) | 0.056 |
| Psychoses | 2,327 (12.1%) | 7,433 (7.7%) | 0.147 |
| Depression | 3,617 (18.8%) | 11,856 (12.3%) | 0.179 |
| Lives in an MSA | 16,726 (87.0%) | 81,245 (84.5%) | 0.071 |
| Possible ALS Symptoms |  |  |  |
| Any | 16,097 (83.7%) | 48,713 (50.7%) | 0.752 |
| By Origin |  |  |  |
| Bulbar | 5,561 (28.9%) | 7,679 (8.0%) | 0.560 |
| Motor | 15,208 (79.1%) | 47,089 (49.0%) | 0.661 |
| By Number of Major Classes |  |  |  |
| No Symptoms | 3,129 (16.3%) | 47,413 (49.3%) | 0.831 |
| Only Motor or Only Bulbar | 11,425 (59.4%) | 42,658 (44.4%) |  |
| Both Motor and Bulbar | 4,672 (24.3%) | 6,055 (6.3%) |  |
| By Symptom Class |  |  |  |
| Speech Problems | 5,356 (27.9%) | 6,620 (6.9%) | 0.576 |
| Swallowing Problems | 572 (3.0%) | 1,465 (1.5%) | 0.098 |
| Muscle Problems | 6,749 (35.1%) | 5,028 (5.2%) | 0.802 |
| Gait Problems | 5,761 (30.0%) | 8,373 (8.7%) | 0.559 |
| Involuntary Movements | 4,561 (23.7%) | 6,078 (6.3%) | 0.502 |
| Pain | 11,147 (58.0%) | 41,847 (43.5%) | 0.292 |
| Falls | 1,142 (5.9%) | 2,567 (2.7%) | 0.162 |
| Other Motor | 9,050 (47.1%) | 14,195 (14.8%) | 0.746 |
| By Number of Symptoms |  |  |  |
| 0 | 3,129 (16.3%) | 47,413 (49.3%) | 1.008 |
| 1 | 4,254 (22.1%) | 28,525 (29.7%) |  |
| 2 | 3,481 (18.1%) | 10,312 (10.7%) |  |
| 3 | 3,451 (17.9%) | 5,302 (5.5%) |  |
| 4 | 2,663 (13.9%) | 2,596 (2.7%) |  |
| 5 | 1,523 (7.9%) | 1,296 (1.3%) |  |
| 6 | 578 (3.0%) | 541 (0.6%) |  |
| 7 | 142 (0.7%) | 134 (0.1%) |  |
| 8 | <10 (0.0%) | <10 (0.0%) |  |

Both having one or more ALS symptoms (OR = 4.92; 95% CI: 4.36, 5.56) and living in an urban area (OR = 1.44; 95% CI: 1.28, 1.61) were independent risk factors for getting a diagnosis of ALS, **Table 3**. The interaction between having an ALS symptom and living in an urban area is less than 1 (OR = 0.83; 95% CI: 0.73, 0.94). In other words, having a symptom in a rural dweller increases the odds of an ALS diagnosis by a factor of 4.92 while the same symptom has a smaller increase in the odds when the person lives in an urban area (0.83 x 4.92 = 4.08). A simple way to understand this result is that having an ALS-like symptom partially reduces the urban bias in ALS diagnosis. Considering only motor or only bulbar symptoms had similar increases in the odds of getting an ALS diagnosis (OR = 3.75 and 3.80, respectively). The odds ratio for living in an urban area and the interactive effect was similar between all three models. Full model details are included in supplemental **Tables S1-S3**.

**Table 3:** Adjusted odds ratios for acquiring a future diagnosis of ALS after having any of the study symptoms, only bulbar, or only motor symptoms, living in and urban area, and the interaction of having the symptom and living in an urban area.

|  | Having any ALS Symptoms | Having Only Bulbar Symptoms | Having Only Motor Symptoms |
| --- | --- | --- | --- |
| Has Symptom | 4.92 (4.36, 5.56) | 3.80 (3.37, 4.29) | 3.75 (3.35, 4.19) |
| Lives in Urban Area | 1.44 (1.28, 1.61) | 1.27 (1.20, 1.35) | 1.43 (1.29, 1.58) |
| Has Symptom and Lives in Urban Area | 0.83 (0.73, 0.94) | 0.84 (0.74, 0.95) | 0.82 (0.73, 0.92) |

The individual symptoms had odds ratios above 1, **Figure 1**, and all except falls and swallowing problems were statistically significant. The main effect for living in a metropolitan county was similar at an odds ratio of approximately 1.3 across all 8 symptom-specific models. The interactive effect was significant and negative for 4 symptoms (speech problems, muscle weakness, pain, and other symptoms), non-significant and negative for two (gait problems, involuntary movements) and non-significant and positive for two (swallowing problems, falls). The full set of coefficients are reported in **Tables S4-S11**.

**Figure 1:**
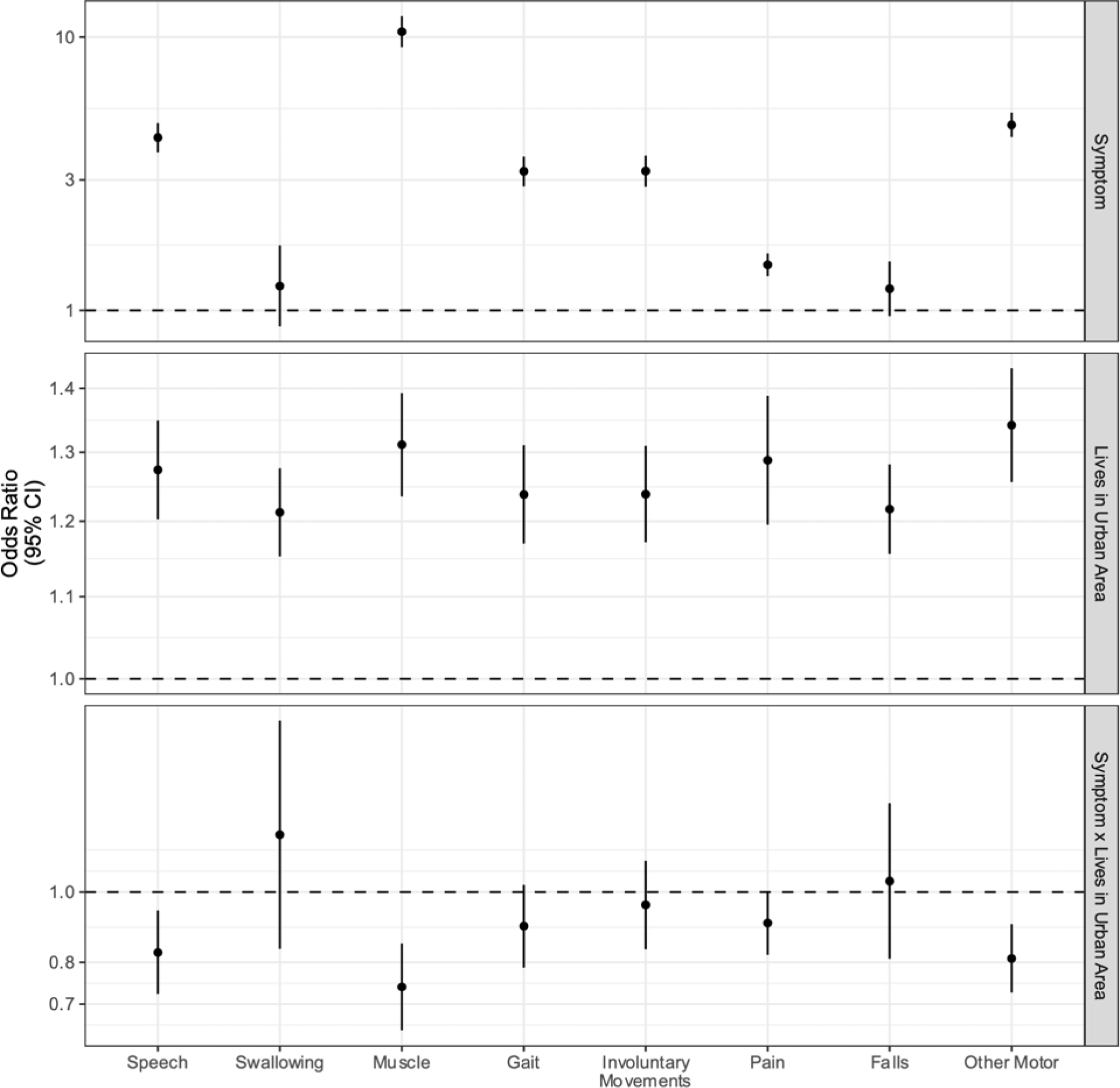
Odds Ratio for a Future Diagnosis of ALS by Specific Symptom. All symptoms were associated with increased odds of a future ALS diagnosis; however, the effect was variable by the specific symptom. More common and less ALS-specific symptoms had lower odds ratios. Points are the estimated OR, lines denote the 95% CI.

There was a dose-response relationship between the number of symptoms and the odds of developing ALS, **Table 4**. While any number of symptoms was associated with statistically significant increased odds, the greatest increase occurring at 5 symptoms (OR=33.20). While having 6+ symptoms was still associated with increased odds, the odds ratios were lower than the peak at 5 symptoms. The interaction effect between number of symptoms and living in an urban area remained consistent at an OR near 0.7.

**Table 4:** Full model estimates for acquiring an ALS diagnosis by number of symptoms. Note the interaction between living in an urban area and having 8 symptoms could not be estimated (there were not individuals with 8 symptoms in both rural and urban settings).

|  | Odds Ratio | 95% CI |  |
| --- | --- | --- | --- |
|  |  | Lower Bound | Upper Bound |
| *Health Care Utilization History* |  |  |  |
| Number of Outpatient Visits Per Year | 1.02 | 1.01 | 1.02 |
| Number of Inpatient Visits Per year | 0.79 | 0.74 | 0.84 |
| Mean Number of Diagnoses Per Outpatient Visit | 1.06 | 1.02 | 1.10 |
| Number of Inpatient Days Per Year | 0.99 | 0.99 | 1.00 |
| *Elixhauser Comorbidities* |  |  |  |
| CHF | 0.64 | 0.59 | 0.69 |
| Valvular Diseases | 0.96 | 0.90 | 1.02 |
| Pulm. HTN | 0.99 | 0.89 | 1.09 |
| PVD | 0.84 | 0.79 | 0.89 |
| HTN | 0.98 | 0.94 | 1.03 |
| HTN with Complications | 0.90 | 0.84 | 0.96 |
| Paralysis | 2.21 | 2.04 | 2.39 |
| Neurological Disorders | 4.14 | 3.94 | 4.34 |
| COPD | 0.94 | 0.90 | 0.99 |
| Diabetes | 0.89 | 0.84 | 0.94 |
| Diabetes with Complications | 0.77 | 0.71 | 0.84 |
| Hypothyroidism | 1.09 | 1.03 | 1.15 |
| Renal Disease | 0.61 | 0.56 | 0.67 |
| Liver Disease | 0.93 | 0.85 | 1.01 |
| Peptic Ulcer Disease | 0.98 | 0.77 | 1.25 |
| HIV | 1.13 | 0.78 | 1.66 |
| Lymphoma | 1.24 | 1.08 | 1.43 |
| Metastatic Cancer | 0.35 | 0.30 | 0.40 |
| Solid Tumor | 0.87 | 0.82 | 0.93 |
| Rheumatic Disorders | 1.11 | 1.03 | 1.18 |
| Coagulopathy | 0.84 | 0.77 | 0.92 |
| Obesity | 0.78 | 0.73 | 0.83 |
| Weight Loss | 1.55 | 1.45 | 1.66 |
| Fluid and Electrolytes Disorders | 0.74 | 0.70 | 0.79 |
| Blood Loss | 0.67 | 0.58 | 0.76 |
| Anemia | 1.02 | 0.96 | 1.07 |
| Alcohol Abuse | 0.86 | 0.76 | 0.98 |
| Drug Abuse | 0.80 | 0.69 | 0.92 |
| Psychoses | 0.80 | 0.75 | 0.85 |
| Depression | 0.90 | 0.85 | 0.95 |
| *ALS Symptom History* |  |  |  |
| Never Had Any Symptoms | Reference | --- | --- |
| Had 1 Symptom | 2.63 | 2.28 | 3.04 |
| Had 2 Symptoms | 7.42 | 6.35 | 8.68 |
| Had 3 Symptoms | 15.17 | 12.79 | 17.99 |
| Had 4 Symptoms | 26.65 | 21.72 | 32.70 |
| Had 5 Symptoms | 33.20 | 24.80 | 44.44 |
| Had 6 Symptoms | 22.63 | 14.96 | 34.23 |
| Had 7 Symptoms | 16.36 | 7.78 | 34.41 |
| Had 8 Symptoms | 7.66 | 2.29 | 25.58 |
| *Urban Residence* |  |  |  |
| Lives in Non-Metropolitan County | Reference | --- | --- |
| Lives in Metropolitan County | 1.46 | 1.30 | 1.63 |
| *Interaction Between Residency and Symptoms* |  |  |  |
| Lives in Metropolitan County **and** has 1 Symptom | 0.97 | 0.84 | 1.14 |
| Lives in Metropolitan County **and** has 2 Symptom | 0.79 | 0.67 | 0.93 |
| Lives in Metropolitan County **and** has 3 Symptom | 0.75 | 0.63 | 0.90 |
| Lives in Metropolitan County **and** has 4 Symptom | 0.73 | 0.59 | 0.91 |
| Lives in Metropolitan County **and** has 5 Symptom | 0.67 | 0.49 | 0.90 |
| Lives in Metropolitan County **and** has 6 Symptom | 0.91 | 0.58 | 1.40 |
| Lives in Metropolitan County **and** has 7 Symptom | 1.38 | 0.62 | 3.06 |
| Lives in Metropolitan County **and** has 8 Symptom | --- | --- | --- |

The odds of an ALS diagnosis following ALS-like symptoms is persistently high even we imposing exclusion periods of over a year, **Figure 2**. Full coefficients are reported in **Table S12** for selected delays. Starting approximately 1 year before the diagnosis date, the relationship between having ALS-like symptoms and a future ALS diagnosis starts to increase rapidly. This increase in the odds ratio indicates an increase in ALS-like symptoms starting at least a year prior to the ALS diagnosis date.

**Figure 2:**
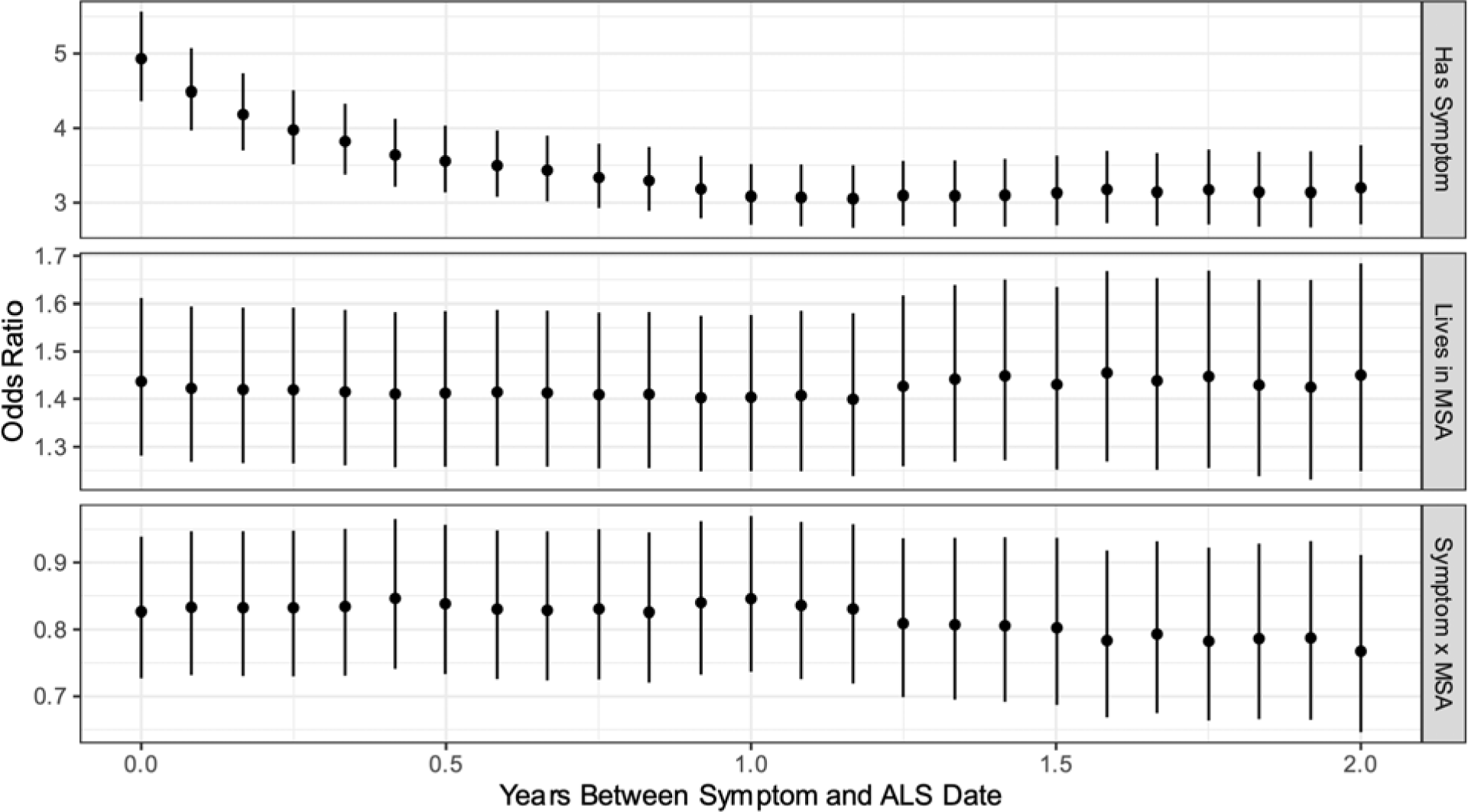
Odds Ratio for a Future Diagnosis of ALS by Exclusion Period Length. For each exclusion period, we estimate a model where we exclude anyone who had their first ALS-like symptoms within the exclusion period before their ALS diagnosis. For example, if the exclusion period was 90 days, we would remove any cases of ALS where the first time they were diagnosed with the ALS-like symptom was less than 90 days before they were diagnosed with ALS. Each point is the estimate from a model incorporating that exclusion period and the lines denote 95% CIs. The OR associated with ALS-like symptoms starts to increase rapidly starting approximately 1 year before the person is diagnosed with ALS. This suggests ALS symptoms may start appearing and being noted clinically at least 1 year before the diagnosis of ALS is made.

Using the Kaplan-Meier estimator, we find a younger age at diagnosis of ALS for those who live in urban areas compared to those who do not, **Figure 3A**. Having any symptoms, **Figure 3B**, and having more symptoms, **Figure 3C**, also increases the hazard of developing ALS. When considered jointly, **Figure 3D**, both having symptoms and living in a metropolitan area increase the hazard of being diagnosed with ALS; however, the difference between the urban and rural Kaplan-Meier curves is smaller in those with symptoms than without. Cox proportional hazards regression shows that these shifts are statistically significant and persist after adjustment for confounders, **Table 5**.

**Figure 3:**
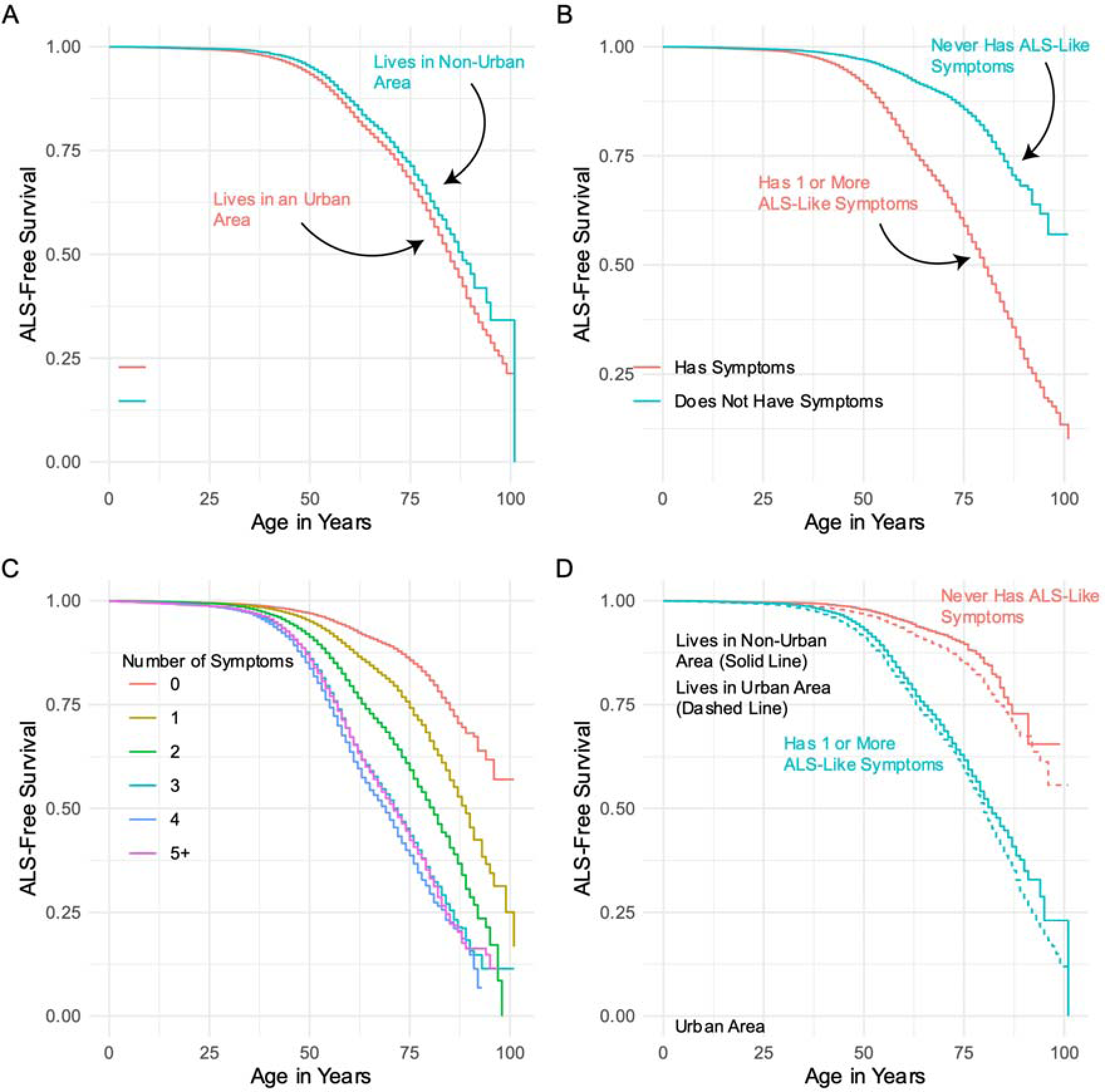
Kaplan-Meier Survival Curves for Age at ALS Diagnosis. Panel A shows a younger age-at-diagnosis among people living in urban vs non-urban locations. Panel B shows a younger age-at-diagnosis for people with vs without ALS-like symptoms. Panel C shows younger age-at-diagnosis with more ALS-like symptoms. Panel D shows an interaction between having symptoms and living in an urban area – both with and without symptoms, people in urban areas are diagnosed at younger ages than their non-urban peers; however, the difference between urban and non-urban patients with a symptom history is much smaller.

**Table 5:** Cox Proportional Hazards Regression on Age at Diagnosis with ALS. Hazard ratios > 1 indicate younger age at diagnosis.

|  |  | 95% CI |  |
| --- | --- | --- | --- |
|  | Hazard Ratio | Lower Bound | Upper Bound |
| *Health Care Utilization History* |  |  |  |
| Number of Outpatient Visits Per Year | 1.02 | 1.01 | 1.02 |
| Number of Inpatient Visits Per year | 0.82 | 0.78 | 0.86 |
| Mean Number of Diagnoses Per Outpatient Visit | 1.12 | 1.09 | 1.16 |
| Number of Inpatient Days Per Year | 0.99 | 0.99 | 1.00 |
| *Elixhauser Comorbidities* |  |  |  |
| CHF | 0.69 | 0.65 | 0.75 |
| Valvular Diseases | 0.98 | 0.93 | 1.03 |
| Pulm. HTN | 0.99 | 0.91 | 1.09 |
| PVD | 0.94 | 0.89 | 0.99 |
| HTN | 0.99 | 0.95 | 1.03 |
| HTN with Complications | 0.92 | 0.87 | 0.98 |
| Paralysis | 2.17 | 2.05 | 2.30 |
| Neurological Disorders | 4.08 | 3.93 | 4.24 |
| COPD | 0.98 | 0.94 | 1.01 |
| Diabetes | 0.88 | 0.84 | 0.93 |
| Diabetes with Complications | 0.84 | 0.78 | 0.90 |
| Hypothyroidism | 1.10 | 1.05 | 1.15 |
| Renal Disease | 0.65 | 0.59 | 0.70 |
| Liver Disease | 0.96 | 0.90 | 1.03 |
| Peptic Ulcer Disease | 1.04 | 0.84 | 1.28 |
| HIV | 0.99 | 0.71 | 1.38 |
| Lymphoma | 1.18 | 1.05 | 1.33 |
| Metastatic Cancer | 0.39 | 0.34 | 0.44 |
| Solid Tumor | 0.90 | 0.85 | 0.95 |
| Rheumatic Disorders | 1.22 | 1.16 | 1.29 |
| Coagulopathy | 0.88 | 0.81 | 0.96 |
| Obesity | 0.85 | 0.81 | 0.90 |
| Weight Loss | 1.54 | 1.45 | 1.62 |
| Fluid and Electrolytes Disorders | 0.84 | 0.80 | 0.89 |
| Blood Loss | 0.75 | 0.66 | 0.84 |
| Anemia | 1.04 | 1.00 | 1.09 |
| Alcohol Abuse | 0.85 | 0.77 | 0.95 |
| Drug Abuse | 0.85 | 0.76 | 0.96 |
| Psychoses | 0.86 | 0.82 | 0.91 |
| Depression | 0.98 | 0.94 | 1.03 |
| *ALS Symptom History* |  |  |  |
| Never Had Any Symptoms | Reference | --- | --- |
| Had 1+ Symptoms | 4.67 | 4.17 | 5.24 |
| *Urban Residence* |  |  |  |
| Lives in Non-Metropolitan County | Reference | --- | --- |
| Lives in Metropolitan County | 1.42 | 1.27 | 1.58 |
| *Interaction Between Residency and Symptoms* |  |  |  |
| Lives in Metropolitan County **and** has Symptoms | 0.82 | 0.73 | 0.92 |

## Discussion

We tested the hypotheses that 1) symptoms of ALS appear in administrative record before the ALS diagnosis, 2) there are urban/rural differences in the rate of diagnosis, and 3) there are interactions between these exposures. We found having any of eight ALS symptoms is associated with a nearly 5-fold increase in the odds of being diagnosed with ALS in the future. Across all models, urban dwellers had greater odds of being diagnosed with ALS; however, the effect of having a symptom was smaller among urban residents compared to rural residents.

We hypothesized that ALS diagnosis was more common in urban areas due to greater access to care. This is supported by the finding of interaction odds ratios less than 1 (symptoms matter less for future diagnosis in urban versus non-urban settings). Additionally, our survival analysis found that having symptoms or living in an urban area was associated with a younger age at ALS diagnosis. If the age-dependent incidence of ALS is similar between urban and rural places, (13, 14) this finding would suggest people with ALS are being diagnosed earlier in their disease course and with presumably lower symptom burden when they live in urban areas.

Indeed, there is limited evidence suggesting that ALS may be more common in rural settings because of greater pesticide exposures.(13, 15) If this is the case, our result would be stronger as we found a greater incidence of diagnosis among urban dwellers.

There appeared to be no difference in whether these were chiefly motor or bulbar symptoms; both were associated with a nearly 4-fold increase in the odds of developing an ALS diagnosis. Certain symptoms had greater impacts on the odds of acquiring an ALS diagnosis. For instance, muscle weakness had an OR of nearly 10 while other, less specific, or more common symptoms (e.g., falls) had lower ORs. There was a dose-response pattern between the number of symptoms and the odds of being diagnosed with ALS which peaked at five symptoms (OR = 33.20 compared to a person without symptoms). The decline in ORs at very high number of symptoms likely stems from ALS having been considered and ruled out for people with many ALS-like symptoms but without a diagnosis of ALS.

Our finding that ALS-like symptoms predate the diagnosis of ALS is supported by prior literature. A chart review of people diagnosed with ALS found the earliest symptoms included limb and mobility problems, bulbar symptoms, and respiratory problems.(16) Another study of 202 people with limb-onset ALS found that half of patients first presented to a non-neurologist, which had implications for time to diagnosis – patients who presented to orthopedists had a longer time to diagnosis.(17) Our work directly builds and expands upon the prior analysis performed using Medicare claims finding increased rates of the 8 symptoms considered here.(7) Our findings and the broader literature are consistent with current paradigms for prodromal and other early ALS symptoms.

Our study has several limitations. First, inherent to all retrospective database studies, we are limited by what is recorded in the administrative record. We observe symptoms only through diagnoses and only observe diagnoses that are recorded. This is necessarily a less rich data source than contained in a chart or clinical evaluation. Second, the presentation of ALS may overlap with other neurological diseases and up to 10% of people with ALS may be undiagnosed.(18, 19). While the ALS diagnostic codes are likely very specific (>99%) with responsible sensitivity (78.9%-91.6%) these codes may follow a series of delayed or inaccurate non-ALS diagnoses.(20, 21, 22) Third, we classified urban as having resided in a metropolitan county. This may misclassify some residents of rural parts of counties as urban. Additionally, an MSA is a metro area of at least 50,000 people: a city of 50,000 people and of 1,000,000 are treated the same. Within the classification, there exists significant heterogeneity in terms of access to specialist health care within that case.

In both rural and urban settings, retrospective analyses consistently find a year or longer period before diagnosis when symptoms are present. There is an urgent and unmet need to accelerate the detection and diagnostic timeline for people living with ALS. Our results suggest that residents of rural communities, who are disproportionately elderly and face limited access to specialist care, are more dependent on major symptoms to lead to a diagnosis of ALS compared to urban residents. With later and more severe disease at diagnosis, these individuals will have spent more time living with the unmitigated symptoms of ALS and have higher risk of falls, injuries, and other preventable morbidity. Educational outreach and telehealth have the potential to reduce some of these disparate outcomes (23, 24); however, any effort to improve the detection-to-diagnosis process will need to consider the particular needs of rural populations. A failure of improvement to consider the barriers and challenges of the rural elderly will exacerbate the existing disparities in access to care, diagnosis, and outcomes.

## Data Availability and Deposition

The data used in this analysis was used under a license agreement with Merative Marketscan. The authors cannot redistribute this data; however, interested users may contact Merative for licensing terms.

## Code Availability

The data extraction, processing, and analysis code is shared at <github.com>

