## Supplemental Tables for "Rurality Modifies the Association Between Symptoms and the Diagnosis of Amyotrophic Lateral Sclerosis"

Table S1: Full model estimates for acquiring an ALS diagnosis following having any ALS symptoms.

|  |  | 95% CI | |
| --- | --- | --- | --- |
|  | Odds Ratio | Lower Bound | Upper Bound |
| *Health Care Utilization History* |  |  |  |
| Number of Outpatient Visits Per Year | 1.02 | 1.02 | 1.03 |
| Number of Inpatient Visits Per year | 0.78 | 0.84 | 0.82 |
| Mean Number of Diagnoses Per Outpatient Visit | 1.15 | 1.10 | 1.19 |
| Number of Inpatient Days Per Year | 0.99 | 0.99 | 1.00 |
| *Elixhauser Comorbidities* |  |  |  |
| CHF | 0.64 | 0.59 | 0.69 |
| Valvular Diseases | 0.97 | 0.92 | 1.03 |
| Pulm. HTN | 0.99 | 0.90 | 1.09 |
| PVD | 0.91 | 0.86 | 0.96 |
| HTN | 0.98 | 0.94 | 1.02 |
| HTN with Complications | 0.91 | 0.85 | 0.97 |
| Paralysis | 3.00 | 2.78 | 3.23 |
| Neurological Disorders | 5.21 | 4.98 | 5.45 |
| COPD | 0.95 | 0.91 | 1.00 |
| Diabetes | 0.86 | 0.82 | 0.91 |
| Diabetes with Complications | 0.80 | 0.74 | 0.86 |
| Hypothyroidism | 1.11 | 1.05 | 1.16 |
| Renal Disease | 0.60 | 0.55 | 0.65 |
| Liver Disease | 0.94 | 0.87 | 1.02 |
| Peptic Ulcer Disease | 1.00 | 0.80 | 1.25 |
| HIV | 0.98 | 0.68 | 1.40 |
| Lymphoma | 1.20 | 1.05 | 1.38 |
| Metastatic Cancer | 0.33 | 0.29 | 0.38 |
| Solid Tumor | 0.86 | 0.81 | 0.92 |
| Rheumatic Disorders | 1.24 | 1.17 | 1.32 |
| Coagulopathy | 0.85 | 0.78 | 0.93 |
| Obesity | 0.83 | 0.78 | 0.88 |
| Weight Loss | 1.69 | 1.59 | 1.81 |
| Fluid and Electrolytes Disorders | 0.80 | 0.75 | 0.85 |
| Blood Loss | 0.70 | 0.61 | 0.79 |
| Anemia | 1.05 | 0.99 | 1.10 |
| Alcohol Abuse | 0.83 | 0.74 | 0.93 |
| Drug Abuse | 0.83 | 0.72 | 0.94 |
| Psychoses | 0.82 | 0.77 | 0.87 |
| Depression | 0.97 | 0.92 | 1.02 |
| *ALS Symptom History* |  |  |  |
| Never Had ALS Symptoms | Reference | --- | --- |
| Had 1+ ALS Symptoms | 4.92 | 4.36 | 5.56 |
| *Urban Residence* |  |  |  |
| Lives in Non-Metropolitan County | Reference | --- | --- |
| Lives in Metropolitan County | 1.44 | 1.28 | 1.61 |
| *Interaction Between Residency and Symptoms* |  |  |  |
| Lives in Metropolitan County **and** has ALS Symptoms | 0.83 | 0.73 | 0.94 |

Table S2: Full model estimates for acquiring an ALS diagnosis following bulbar symptoms.

|  |  | 95% CI | |
| --- | --- | --- | --- |
|  | Odds Ratio | Lower Bound | Upper Bound |
| *Health Care Utilization History* |  |  |  |
| Number of Outpatient Visits Per Year | 1.03 | 1.03 | 1.03 |
| Number of Inpatient Visits Per year | 0.78 | 0.74 | 0.83 |
| Mean Number of Diagnoses Per Outpatient Visit | 1.29 | 1.25 | 1.34 |
| Number of Inpatient Days Per Year | 0.99 | 0.99 | 1.00 |
| *Elixhauser Comorbidities* |  |  |  |
| CHF | 0.60 | 0.55 | 0.65 |
| Valvular Diseases | 0.94 | 0.89 | 1.00 |
| Pulm. HTN | 0.94 | 0.85 | 1.04 |
| PVD | 0.92 | 0.87 | 0.98 |
| HTN | 1.03 | 0.99 | 1.08 |
| HTN with Complications | 0.88 | 0.82 | 0.94 |
| Paralysis | 3.00 | 2.77 | 3.24 |
| Neurological Disorders | 5.23 | 4.99 | 5.47 |
| COPD | 0.93 | 0.89 | 0.97 |
| Diabetes | 0.85 | 0.80 | 0.90 |
| Diabetes with Complications | 0.80 | 0.74 | 0.87 |
| Hypothyroidism | 1.10 | 1.04 | 1.15 |
| Renal Disease | 0.55 | 0.50 | 0.60 |
| Liver Disease | 0.95 | 0.88 | 1.03 |
| Peptic Ulcer Disease | 0.92 | 0.72 | 1.16 |
| HIV | 0.85 | 0.59 | 1.22 |
| Lymphoma | 1.15 | 1.00 | 1.32 |
| Metastatic Cancer | 0.30 | 0.26 | 0.34 |
| Solid Tumor | 0.84 | 0.79 | 0.89 |
| Rheumatic Disorders | 1.36 | 1.27 | 1.45 |
| Coagulopathy | 0.83 | 0.76 | 0.91 |
| Obesity | 0.84 | 0.79 | 0.90 |
| Weight Loss | 1.57 | 1.47 | 1.67 |
| Fluid and Electrolytes Disorders | 0.77 | 0.73 | 0.82 |
| Blood Loss | 0.65 | 0.57 | 0.74 |
| Anemia | 1.05 | 1.00 | 1.11 |
| Alcohol Abuse | 0.88 | 0.78 | 0.99 |
| Drug Abuse | 0.83 | 0.72 | 0.96 |
| Psychoses | 0.79 | 0.74 | 0.84 |
| Depression | 0.96 | 0.91 | 1.02 |
| *ALS Symptom History* |  |  |  |
| Never Had Bulbar Symptoms | Reference | --- | --- |
| Had 1+ Bulbar Symptoms | 3.80 | 3.37 | 4.29 |
| *Urban Residence* |  |  |  |
| Lives in Non-Metropolitan County | Reference | --- | --- |
| Lives in Metropolitan County | 1.27 | 1.20 | 1.35 |
| *Interaction Between Residency and Symptoms* |  |  |  |
| Lives in Metropolitan County **and** has Bulbar Symptoms | 0.84 | 0.74 | 0.95 |

Table S3: Full model estimates for acquiring an ALS diagnosis following motor symptoms.

|  |  | 95% CI | |
| --- | --- | --- | --- |
|  | Odds Ratio | Lower Bound | Upper Bound |
| *Health Care Utilization History* |  |  |  |
| Number of Outpatient Visits Per Year | 1.03 | 1.02 | 1.03 |
| Number of Inpatient Visits Per year | 0.78 | 0.74 | 0.82 |
| Mean Number of Diagnoses Per Outpatient Visit | 1.19 | 1.15 | 1.23 |
| Number of Inpatient Days Per Year | 1.00 | 0.99 | 1.00 |
| *Elixhauser Comorbidities* |  |  |  |
| CHF | 0.63 | 0.58 | 0.68 |
| Valvular Diseases | 0.97 | 0.82 | 1.02 |
| Pulm. HTN | 0.98 | 0.89 | 1.08 |
| PVD | 0.91 | 0.86 | 0.96 |
| HTN | 0.98 | 0.94 | 1.02 |
| HTN with Complications | 0.91 | 0.85 | 0.96 |
| Paralysis | 3.03 | 2.81 | 3.27 |
| Neurological Disorders | 5.42 | 5.18 | 5.67 |
| COPD | 0.97 | 0.93 | 1.01 |
| Diabetes | 0.86 | 0.81 | 0.90 |
| Diabetes with Complications | 0.79 | 0.73 | 0.85 |
| Hypothyroidism | 1.12 | 1.06 | 1.17 |
| Renal Disease | 0.59 | 0.54 | 0.64 |
| Liver Disease | 0.94 | 0.87 | 1.02 |
| Peptic Ulcer Disease | 1.02 | 0.82 | 1.28 |
| HIV | 0.99 | 0.69 | 1.41 |
| Lymphoma | 1.19 | 1.04 | 1.36 |
| Metastatic Cancer | 0.33 | 0.29 | 0.37 |
| Solid Tumor | 0.86 | 0.81 | 0.91 |
| Rheumatic Disorders | 1.25 | 1.17 | 1.33 |
| Coagulopathy | 0.84 | 0.77 | 0.92 |
| Obesity | 0.83 | 0.78 | 0.88 |
| Weight Loss | 1.73 | 1.62 | 1.84 |
| Fluid and Electrolytes Disorders | 0.80 | 0.75 | 0.84 |
| Blood Loss | 0.69 | 0.61 | 0.79 |
| Anemia | 1.04 | 0.99 | 1.09 |
| Alcohol Abuse | 0.83 | 0.74 | 0.94 |
| Drug Abuse | 0.81 | 0.71 | 0.93 |
| Psychoses | 0.81 | 0.76 | 0.86 |
| Depression | 0.97 | 0.92 | 1.02 |
| *ALS Symptom History* |  |  |  |
| Never Had Motor Symptoms | Reference | --- | --- |
| Had 1+ Motor Symptoms | 3.75 | 3.35 | 4.19 |
| *Urban Residence* |  |  |  |
| Lives in Non-Metropolitan County | Reference | --- | --- |
| Lives in Metropolitan County | 1.43 | 1.29 | 1.58 |
| *Interaction Between Residency and Symptoms* |  |  |  |
| Lives in Metropolitan County **and** has Motor Symptoms | 0.82 | 0.73 | 0.92 |

Table S4: Full model estimates for acquiring an ALS diagnosis following speech symptoms.

|  |  | 95% CI | |
| --- | --- | --- | --- |
|  | Odds Ratio | Lower Bound | Upper Bound |
| *Health Care Utilization History* |  |  |  |
| Number of Outpatient Visits Per Year | 1.03 | 1.03 | 1.03 |
| Number of Inpatient Visits Per year | 0.78 | 0.78 | 0.83 |
| Mean Number of Diagnoses Per Outpatient Visit | 1.29 | 1.25 | 1.34 |
| Number of Inpatient Days Per Year | 0.99 | 0.99 | 1.00 |
| *Elixhauser Comorbidities* |  |  |  |
| CHF | 0.59 | 0.55 | 0.64 |
| Valvular Diseases | 0.94 | 0.89 | 1.00 |
| Pulm. HTN | 0.93 | 0.84 | 1.03 |
| PVD | 0.92 | 0.87 | 0.98 |
| HTN | 1.04 | 0.99 | 1.08 |
| HTN with Complications | 0.88 | 0.83 | 0.94 |
| Paralysis | 2.96 | 2.74 | 3.20 |
| Neurological Disorders | 5.15 | 4.92 | 5.39 |
| COPD | 0.93 | 0.89 | 0.97 |
| Diabetes | 0.85 | 0.80 | 0.90 |
| Diabetes with Complications | 0.80 | 0.74 | 0.87 |
| Hypothyroidism | 1.10 | 1.05 | 1.16 |
| Renal Disease | 0.55 | 0.50 | 0.60 |
| Liver Disease | 0.95 | 0.88 | 1.03 |
| Peptic Ulcer Disease | 0.90 | 0.71 | 1.14 |
| HIV | 0.87 | 0.60 | 1.24 |
| Lymphoma | 1.16 | 1.01 | 1.33 |
| Metastatic Cancer | 0.30 | 0.26 | 0.34 |
| Solid Tumor | 0.84 | 0.79 | 0.89 |
| Rheumatic Disorders | 1.36 | 1.28 | 1.45 |
| Coagulopathy | 0.83 | 0.76 | 0.91 |
| Obesity | 0.85 | 0.80 | 0.90 |
| Weight Loss | 1.55 | 1.45 | 1.66 |
| Fluid and Electrolytes Disorders | 0.77 | 0.73 | 0.82 |
| Blood Loss | 0.65 | 0.57 | 0.74 |
| Anemia | 1.05 | 1.00 | 1.11 |
| Alcohol Abuse | 0.87 | 0.77 | 0.98 |
| Drug Abuse | 0.83 | 0.72 | 0.96 |
| Psychoses | 0.79 | 0.74 | 0.84 |
| Depression | 0.96 | 0.91 | 1.02 |
| *ALS Symptom History* |  |  |  |
| Never Had Speech Symptoms | Reference | --- | --- |
| Had 1+ Speech Symptoms | 4.28 | 3.78 | 4.85 |
| *Urban Residence* |  |  |  |
| Lives in Non-Metropolitan County | Reference | --- | --- |
| Lives in Metropolitan County | 1.27 | 1.20 | 1.35 |
| *Interaction Between Residency and Symptoms* |  |  |  |
| Lives in Metropolitan County **and** has Speech Symptoms | 0.83 | 0.72 | 0.94 |

Table S5: Full model estimates for acquiring an ALS diagnosis following swallowing symptoms.

|  |  | 95% CI | |
| --- | --- | --- | --- |
|  | Odds Ratio | Lower Bound | Upper Bound |
| *Health Care Utilization History* |  |  |  |
| Number of Outpatient Visits Per Year | 1.03 | 1.03 | 1.04 |
| Number of Inpatient Visits Per year | 0.77 | 0.73 | 0.82 |
| Mean Number of Diagnoses Per Outpatient Visit | 1.33 | 1.29 | 1.37 |
| Number of Inpatient Days Per Year | 1.00 | 0.99 | 1.00 |
| *Elixhauser Comorbidities* |  |  |  |
| CHF | 0.60 | 0.56 | 0.65 |
| Valvular Diseases | 0.96 | 0.91 | 1.02 |
| Pulm. HTN | 0.95 | 0.86 | 1.05 |
| PVD | 0.94 | 0.88 | 0.99 |
| HTN | 1.03 | 0.99 | 1.07 |
| HTN with Complications | 0.89 | 0.84 | 0.95 |
| Paralysis | 3.30 | 3.06 | 3.56 |
| Neurological Disorders | 5.99 | 5.73 | 6.26 |
| COPD | 0.98 | 0.94 | 1.03 |
| Diabetes | 0.85 | 0.80 | 0.90 |
| Diabetes with Complications | 0.78 | 0.73 | 0.85 |
| Hypothyroidism | 1.13 | 1.07 | 1.18 |
| Renal Disease | 0.54 | 0.50 | 0.59 |
| Liver Disease | 0.95 | 0.88 | 1.03 |
| Peptic Ulcer Disease | 1.01 | 0.80 | 1.26 |
| HIV | 0.85 | 0.59 | 1.21 |
| Lymphoma | 1.16 | 1.01 | 1.33 |
| Metastatic Cancer | 0.30 | 0.27 | 0.34 |
| Solid Tumor | 0.84 | 0.80 | 0.90 |
| Rheumatic Disorders | 1.37 | 1.29 | 1.46 |
| Coagulopathy | 0.83 | 0.76 | 0.91 |
| Obesity | 1.75 | 1.64 | 1.86 |
| Weight Loss | 0.80 | 0.75 | 0.84 |
| Fluid and Electrolytes Disorders | 0.67 | 0.58 | 0.76 |
| Blood Loss | 1.05 | 1.00 | 1.10 |
| Anemia | 0.86 | 0.76 | 0.97 |
| Alcohol Abuse | 0.80 | 0.70 | 0.92 |
| Drug Abuse | 1.00 | 0.95 | 1.05 |
| Psychoses | 1.23 | 0.87 | 1.73 |
| Depression | 1.00 | 0.95 | 1.05 |
| *ALS Symptom History* |  |  |  |
| Never Had Swallowing Symptoms | Reference | --- | --- |
| Had 1+ Swallowing Symptoms | 1.23 | 0.87 | 1.73 |
| *Urban Residence* |  |  |  |
| Lives in Non-Metropolitan County | Reference | --- | --- |
| Lives in Metropolitan County | 1.21 | 1.15 | 1.28 |
| *Interaction Between Residency and Symptoms* |  |  |  |
| Lives in Metropolitan County **and** has Swallowing Symptoms | 1.20 | 0.84 | 1.72 |

Table S6: Full model estimates for acquiring an ALS diagnosis following muscle symptoms.

|  |  | 95% CI | |
| --- | --- | --- | --- |
|  | Odds Ratio | Lower Bound | Upper Bound |
| *Health Care Utilization History* |  |  |  |
| Number of Outpatient Visits Per Year | 1.03 | 1.03 | 1.03 |
| Number of Inpatient Visits Per year | 0.77 | 0.72 | 0.81 |
| Mean Number of Diagnoses Per Outpatient Visit | 1.22 | 1.17 | 1.26 |
| Number of Inpatient Days Per Year | 0.99 | 0.99 | 1.00 |
| *Elixhauser Comorbidities* |  |  |  |
| CHF | 0.57 | 0.53 | 0.62 |
| Valvular Diseases | 0.98 | 0.92 | 1.03 |
| Pulm. HTN | 0.91 | 0.82 | 1.02 |
| PVD | 0.88 | 0.82 | 0.93 |
| HTN | 1.05 | 1.00 | 1.09 |
| HTN with Complications | 0.91 | 0.85 | 0.97 |
| Paralysis | 2.33 | 2.14 | 2.53 |
| Neurological Disorders | 5.05 | 4.82 | 5.30 |
| COPD | 1.00 | 0.96 | 1.05 |
| Diabetes | 0.87 | 0.82 | 0.92 |
| Diabetes with Complications | 0.75 | 0.69 | 0.81 |
| Hypothyroidism | 1.14 | 1.08 | 1.20 |
| Renal Disease | 0.52 | 0.47 | 0.57 |
| Liver Disease | 0.97 | 0.89 | 1.05 |
| Peptic Ulcer Disease | 1.06 | 0.83 | 1.36 |
| HIV | 0.94 | 0.64 | 1.38 |
| Lymphoma | 1.17 | 1.01 | 1.35 |
| Metastatic Cancer | 0.30 | 0.26 | 0.35 |
| Solid Tumor | 0.86 | 0.80 | 0.91 |
| Rheumatic Disorders | 1.28 | 1.19 | 1.36 |
| Coagulopathy | 0.80 | 0.73 | 0.88 |
| Obesity | 0.83 | 0.78 | 0.89 |
| Weight Loss | 1.61 | 1.51 | 1.73 |
| Fluid and Electrolytes Disorders | 0.75 | 0.70 | 0.80 |
| Blood Loss | 0.61 | 0.53 | 0.70 |
| Anemia | 1.03 | 0.98 | 1.09 |
| Alcohol Abuse | 0.88 | 0.77 | 1.00 |
| Drug Abuse | 0.83 | 0.72 | 0.97 |
| Psychoses | 0.81 | 0.76 | 0.87 |
| Depression | 0.95 | 0.90 | 1.01 |
| *ALS Symptom History* |  |  |  |
| Never Had Muscle Symptoms | Reference | --- | --- |
| Had 1+ Muscle Symptoms | 10.47 | 9.18 | 11.93 |
| *Urban Residence* |  |  |  |
| Lives in Non-Metropolitan County | Reference | --- | --- |
| Lives in Metropolitan County | 1.31 | 1.24 | 1.39 |
| *Interaction Between Residency and Symptoms* |  |  |  |
| Lives in Metropolitan County **and** has Muscle Symptoms | 0.74 | 0.64 | 0.85 |

Table S7: Full model estimates for acquiring an ALS diagnosis following gait symptoms.

|  |  | 95% CI | |
| --- | --- | --- | --- |
|  | Odds Ratio | Lower Bound | Upper Bound |
| *Health Care Utilization History* |  |  |  |
| Number of Outpatient Visits Per Year | 1.03 | 1.03 | 1.03 |
| Number of Inpatient Visits Per year | 0.74 | 0.70 | 0.79 |
| Mean Number of Diagnoses Per Outpatient Visit | 1.29 | 1.25 | 1.34 |
| Number of Inpatient Days Per Year | 0.99 | 0.99 | 1.00 |
| *Elixhauser Comorbidities* |  |  |  |
| CHF | 0.59 | 0.55 | 0.64 |
| Valvular Diseases | 0.97 | 0.92 | 1.03 |
| Pulm. HTN | 0.92 | 0.83 | 1.02 |
| PVD | 0.89 | 0.84 | 0.94 |
| HTN | 1.02 | 0.98 | 1.07 |
| HTN with Complications | 0.88 | 0.83 | 0.94 |
| Paralysis | 2.69 | 2.48 | 2.91 |
| Neurological Disorders | 5.20 | 4.97 | 5.44 |
| COPD | 1.00 | 0.96 | 1.05 |
| Diabetes | 0.86 | 0.82 | 0.91 |
| Diabetes with Complications | 0.76 | 0.71 | 0.83 |
| Hypothyroidism | 1.14 | 1.08 | 1.20 |
| Renal Disease | 0.55 | 0.50 | 0.60 |
| Liver Disease | 0.97 | 0.90 | 1.05 |
| Peptic Ulcer Disease | 1.06 | 0.84 | 1.33 |
| HIV | 0.87 | 0.61 | 1.25 |
| Lymphoma | 1.18 | 1.03 | 1.36 |
| Metastatic Cancer | 0.30 | 0.27 | 0.35 |
| Solid Tumor | 0.85 | 0.80 | 0.91 |
| Rheumatic Disorders | 1.32 | 1.24 | 1.41 |
| Coagulopathy | 0.82 | 0.75 | 0.89 |
| Obesity | 0.83 | 0.78 | 0.88 |
| Weight Loss | 1.72 | 1.61 | 1.84 |
| Fluid and Electrolytes Disorders | 0.77 | 0.72 | 0.81 |
| Blood Loss | 0.65 | 0.57 | 0.75 |
| Anemia | 1.03 | 0.97 | 1.08 |
| Alcohol Abuse | 0.85 | 0.76 | 0.96 |
| Drug Abuse | 0.80 | 0.70 | 0.93 |
| Psychoses | 0.77 | 0.72 | 0.82 |
| Depression | 0.98 | 0.92 | 1.03 |
| *ALS Symptom History* |  |  |  |
| Never Had Gait Symptoms | Reference | --- | --- |
| Had 1+ Gait Symptoms | 3.22 | 2.84 | 3.65 |
| *Urban Residence* |  |  |  |
| Lives in Non-Metropolitan County | Reference | --- | --- |
| Lives in Metropolitan County | 1.24 | 1.17 | 1.31 |
| *Interaction Between Residency and Symptoms* |  |  |  |
| Lives in Metropolitan County **and** has Gait Symptoms | 0.90 | 0.79 | 1.02 |

Table S8: Full model estimates for acquiring an ALS diagnosis following involuntary movement symptoms.

|  |  | 95% CI | |
| --- | --- | --- | --- |
|  | Odds Ratio | Lower Bound | Upper Bound |
| *Health Care Utilization History* |  |  |  |
| Number of Outpatient Visits Per Year | 1.03 | 1.03 | 1.03 |
| Number of Inpatient Visits Per year | 0.79 | 0.75 | 0.84 |
| Mean Number of Diagnoses Per Outpatient Visit | 1.28 | 1.23 | 1.32 |
| Number of Inpatient Days Per Year | 1.00 | 0.99 | 1.00 |
| *Elixhauser Comorbidities* |  |  |  |
| CHF | 0.61 | 0.57 | 0.66 |
| Valvular Diseases | 0.97 | 0.91 | 1.02 |
| Pulm. HTN | 0.96 | 0.87 | 1.02 |
| PVD | 0.94 | 0.88 | 0.99 |
| HTN | 1.02 | 0.98 | 1.07 |
| HTN with Complications | 0.89 | 0.84 | 0.95 |
| Paralysis | 3.11 | 2.87 | 3.36 |
| Neurological Disorders | 5.57 | 5.33 | 5.83 |
| COPD | 0.97 | 0.93 | 1.02 |
| Diabetes | 0.86 | 0.82 | 0.91 |
| Diabetes with Complications | 0.80 | 0.74 | 0.86 |
| Hypothyroidism | 1.12 | 1.07 | 1.18 |
| Renal Disease | 0.55 | 0.50 | 0.60 |
| Liver Disease | 0.93 | 0.86 | 1.01 |
| Peptic Ulcer Disease | 0.95 | 0.75 | 1.20 |
| HIV | 0.94 | 0.66 | 1.34 |
| Lymphoma | 1.19 | 1.03 | 1.36 |
| Metastatic Cancer | 0.31 | 0.27 | 0.35 |
| Solid Tumor | 0.86 | 0.80 | 0.91 |
| Rheumatic Disorders | 1.30 | 1.22 | 1.39 |
| Coagulopathy | 0.84 | 0.77 | 0.92 |
| Obesity | 0.86 | 0.81 | 0.91 |
| Weight Loss | 1.72 | 1.61 | 1.84 |
| Fluid and Electrolytes Disorders | 0.79 | 0.74 | 0.84 |
| Blood Loss | 0.67 | 0.59 | 0.76 |
| Anemia | 1.05 | 1.00 | 1.10 |
| Alcohol Abuse | 0.85 | 0.76 | 0.96 |
| Drug Abuse | 0.74 | 0.64 | 0.86 |
| Psychoses | 0.78 | 0.73 | 0.84 |
| Depression | 0.97 | 0.92 | 1.02 |
| *ALS Symptom History* |  |  |  |
| Never Had Involuntary Movement Symptoms | Reference | --- | --- |
| Had 1+ Involuntary Movement Symptoms | 3.23 | 2.83 | 3.68 |
| *Urban Residence* |  |  |  |
| Lives in Non-Metropolitan County | Reference | --- | --- |
| Lives in Metropolitan County | 1.24 | 1.17 | 1.31 |
| *Interaction Between Residency and Symptoms* |  |  |  |
| Lives in Metropolitan County **and** has Involuntary Movement Symptoms | 0.96 | 0.83 | 1.10 |

Table S9: Full model estimates for acquiring an ALS diagnosis following pain symptoms.

|  |  | 95% CI | |
| --- | --- | --- | --- |
|  | Odds Ratio | Lower Bound | Upper Bound |
| *Health Care Utilization History* |  |  |  |
| Number of Outpatient Visits Per Year | 1.03 | 1.03 | 1.03 |
| Number of Inpatient Visits Per year | 0.77 | 0.73 | 0.82 |
| Mean Number of Diagnoses Per Outpatient Visit | 1.30 | 1.26 | 1.35 |
| Number of Inpatient Days Per Year | 1.00 | 0.99 | 1.00 |
| *Elixhauser Comorbidities* |  |  |  |
| CHF | 0.61 | 0.56 | 0.65 |
| Valvular Diseases | 0.96 | 0.91 | 1.02 |
| Pulm. HTN | 0.95 | 0.86 | 1.05 |
| PVD | 0.92 | 0.87 | 0.98 |
| HTN | 1.02 | 0.98 | 1.06 |
| HTN with Complications | 0.89 | 0.84 | 0.95 |
| Paralysis | 3.27 | 3.03 | 3.53 |
| Neurological Disorders | 5.97 | 5.71 | 6.24 |
| COPD | 0.98 | 0.93 | 1.02 |
| Diabetes | 0.85 | 0.80 | 0.90 |
| Diabetes with Complications | 0.78 | 0.72 | 0.84 |
| Hypothyroidism | 1.12 | 1.07 | 1.18 |
| Renal Disease | 0.55 | 0.50 | 0.60 |
| Liver Disease | 0.95 | 0.88 | 1.03 |
| Peptic Ulcer Disease | 1.01 | 0.80 | 1.27 |
| HIV | 0.89 | 0.62 | 1.27 |
| Lymphoma | 1.16 | 1.02 | 1.34 |
| Metastatic Cancer | 0.31 | 0.27 | 0.35 |
| Solid Tumor | 0.85 | 0.80 | 0.90 |
| Rheumatic Disorders | 1.32 | 1.24 | 1.41 |
| Coagulopathy | 0.83 | 0.76 | 0.91 |
| Obesity | 0.85 | 0.80 | 0.90 |
| Weight Loss | 1.76 | 1.65 | 1.87 |
| Fluid and Electrolytes Disorders | 0.80 | 0.75 | 0.84 |
| Blood Loss | 0.67 | 0.59 | 0.76 |
| Anemia | 1.04 | 0.99 | 1.10 |
| Alcohol Abuse | 0.85 | 0.76 | 0.96 |
| Drug Abuse | 0.80 | 0.69 | 0.91 |
| Psychoses | 0.80 | 0.75 | 0.85 |
| Depression | 0.99 | 0.94 | 1.04 |
| *ALS Symptom History* |  |  |  |
| Never Had Pain Symptoms | Reference | --- | --- |
| Had 1+ Pain Symptoms | 1.47 | 1.33 | 1.62 |
| *Urban Residence* |  |  |  |
| Lives in Non-Metropolitan County | Reference | --- | --- |
| Lives in Metropolitan County | 1.29 | 1.20 | 1.39 |
| *Interaction Between Residency and Symptoms* |  |  |  |
| Lives in Metropolitan County **and** has Pain Symptoms | 0.91 | 0.82 | 1.00 |

Table S10: Full model estimates for acquiring an ALS diagnosis following fall symptoms.

|  |  | 95% CI | |
| --- | --- | --- | --- |
|  | Odds Ratio | Lower Bound | Upper Bound |
| *Health Care Utilization History* |  |  |  |
| Number of Outpatient Visits Per Year | 1.03 | 1.03 | 1.04 |
| Number of Inpatient Visits Per year | 0.77 | 0.73 | 0.81 |
| Mean Number of Diagnoses Per Outpatient Visit | 1.33 | 1.28 | 1.37 |
| Number of Inpatient Days Per Year | 1.00 | 0.99 | 1.00 |
| *Elixhauser Comorbidities* |  |  |  |
| CHF | 0.60 | 0.56 | 0.65 |
| Valvular Diseases | 0.97 | 0.91 | 1.02 |
| Pulm. HTN | 0.95 | 0.86 | 1.05 |
| PVD | 0.94 | 0.88 | 0.99 |
| HTN | 1.03 | 0.99 | 1.07 |
| HTN with Complications | 0.89 | 0.84 | 0.95 |
| Paralysis | 3.26 | 3.03 | 3.52 |
| Neurological Disorders | 5.96 | 5.70 | 6.23 |
| COPD | 0.98 | 0.94 | 1.03 |
| Diabetes | 0.85 | 0.81 | 0.90 |
| Diabetes with Complications | 0.78 | 0.72 | 0.84 |
| Hypothyroidism | 1.13 | 1.08 | 1.19 |
| Renal Disease | 0.54 | 0.50 | 0.59 |
| Liver Disease | 0.95 | 0.88 | 1.03 |
| Peptic Ulcer Disease | 1.01 | 0.81 | 1.27 |
| HIV | 0.85 | 0.60 | 1.22 |
| Lymphoma | 1.16 | 1.01 | 1.33 |
| Metastatic Cancer | 0.30 | 0.27 | 0.34 |
| Solid Tumor | 0.85 | 0.80 | 0.90 |
| Rheumatic Disorders | 1.37 | 1.29 | 1.46 |
| Coagulopathy | 0.83 | 0.76 | 0.90 |
| Obesity | 0.86 | 0.81 | 0.91 |
| Weight Loss | 1.75 | 1.64 | 1.87 |
| Fluid and Electrolytes Disorders | 0.79 | 0.75 | 0.84 |
| Blood Loss | 0.67 | 0.58 | 0.76 |
| Anemia | 1.05 | 1.00 | 1.10 |
| Alcohol Abuse | 0.85 | 0.76 | 0.96 |
| Drug Abuse | 0.80 | 0.69 | 0.85 |
| Psychoses | 0.79 | 0.75 | 0.85 |
| Depression | 0.99 | 0.94 | 1.05 |
| *ALS Symptom History* |  |  |  |
| Never Had Fall Symptoms | Reference | --- | --- |
| Had 1+ Fall Symptoms | 1.20 | 0.95 | 1.51 |
| *Urban Residence* |  |  |  |
| Lives in Non-Metropolitan County | Reference | --- | --- |
| Lives in Metropolitan County | 1.22 | 1.16 | 1.28 |
| *Interaction Between Residency and Symptoms* |  |  |  |
| Lives in Metropolitan County **and** has Fall Symptoms | 1.04 | 0.81 | 1.33 |

Table S11: Full model estimates for acquiring an ALS diagnosis following other motor symptoms.

|  |  | 95% CI | |
| --- | --- | --- | --- |
|  | Odds Ratio | Lower Bound | Upper Bound |
| *Health Care Utilization History* |  |  |  |
| Number of Outpatient Visits Per Year | 1.03 | 1.02 | 1.03 |
| Number of Inpatient Visits Per year | 0.80 | 0.76 | 0.85 |
| Mean Number of Diagnoses Per Outpatient Visit | 1.23 | 1.19 | 1.27 |
| Number of Inpatient Days Per Year | 1.00 | 0.99 | 1.00 |
| *Elixhauser Comorbidities* |  |  |  |
| CHF | 0.61 | 0.57 | 0.66 |
| Valvular Diseases | 0.96 | 0.91 | 1.02 |
| Pulm. HTN | 0.96 | 0.86 | 1.06 |
| PVD | 0.90 | 0.85 | 0.96 |
| HTN | 1.03 | 0.99 | 1.07 |
| HTN with Complications | 0.90 | 0.84 | 0.96 |
| Paralysis | 2.68 | 2.47 | 2.90 |
| Neurological Disorders | 5.36 | 5.12 | 5.61 |
| COPD | 0.98 | 0.94 | 1.03 |
| Diabetes | 0.86 | 0.82 | 0.91 |
| Diabetes with Complications | 0.77 | 0.71 | 0.83 |
| Hypothyroidism | 1.11 | 1.06 | 1.17 |
| Renal Disease | 0.56 | 0.51 | 0.61 |
| Liver Disease | 0.94 | 0.87 | 1.02 |
| Peptic Ulcer Disease | 0.99 | 0.78 | 1.25 |
| HIV | 0.93 | 0.64 | 1.35 |
| Lymphoma | 1.21 | 1.05 | 1.40 |
| Metastatic Cancer | 0.33 | 0.29 | 0.38 |
| Solid Tumor | 0.86 | 0.81 | 0.92 |
| Rheumatic Disorders | 1.24 | 1.16 | 1.32 |
| Coagulopathy | 0.83 | 0.76 | 0.91 |
| Obesity | 0.82 | 0.77 | 0.87 |
| Weight Loss | 1.69 | 1.59 | 1.81 |
| Fluid and Electrolytes Disorders | 0.77 | 0.72 | 0.81 |
| Blood Loss | 0.65 | 0.57 | 0.75 |
| Anemia | 1.04 | 0.99 | 1.10 |
| Alcohol Abuse | 0.89 | 0.78 | 1.00 |
| Drug Abuse | 0.78 | 0.67 | 0.90 |
| Psychoses | 0.81 | 0.76 | 0.87 |
| Depression | 0.97 | 0.91 | 1.02 |
| *ALS Symptom History* |  |  |  |
| Never Had Other Motor Symptoms | Reference | --- | --- |
| Had 1+ Other Motor Symptoms | 4.77 | 4.31 | 5.28 |
| *Urban Residence* |  |  |  |
| Lives in Non-Metropolitan County | Reference | --- | --- |
| Lives in Metropolitan County | 1.34 | 1.26 | 1.43 |
| *Interaction Between Residency and Symptoms* |  |  |  |
| Lives in Metropolitan County **and** has Other Motor Symptoms | 0.81 | 0.73 | 0.90 |

Table S12: Full model fits with 30, 60, 90, 180, and 365 days of delay.

|  | Delay Length in Days | | | | |
| --- | --- | --- | --- | --- | --- |
|  | 30 | 60 | 90 | 180 | 365 |
| *Health Care Utilization History* |  |  |  |  |  |
| Number of Outpatient Visits Per Year | 1.02 (1.02, 1.03) | 1.03  (1.02, 1.03) | 1.03  (1.03, 1.03) | 1.03  (1.03, 1.03) | 1.03  (1.03, 1.03) |
| Number of Inpatient Visits Per year | 0.78  (0.74, 0.83) | 0.78  (0.73, 0.83) | 0.78  (0.73, 0.83) | 0.78  (0.73, 0.83) | 0.79  (0.73, 0.85) |
| Mean Number of Diagnoses Per Outpatient Visit | 1.14  (1.09, 1.18) | 1.14  (1.10, 1.19) | 1.14  (1.10, 1.19) | 1.13  (1.08, 1.17) | 1.11  (1.06, 1.16) |
| Number of Inpatient Days Per Year | 1.00  (0.99, 1.00) | 1.00  (0.99, 1.00) | 1.00  (0.99, 1.00) | 1.00  (0.99, 1.00) | 1.00  (0.99, 1.00) |
| *Elixhauser Comorbidities* |  |  |  |  |  |
| CHF | 0.63  (0.59, 0.68) | 0.64  (0.59, 0.69) | 0.63  (0.58, 0.68) | 0.63  (0.58, 0.68) | 0.64  (0.59, 0.70) |
| Valvular Diseases | 0.97  (0.92, 1.03) | 0.97  (0.91, 1.02) | 0.97  (0.92, 1.03) | 0.97  0.91, 1.03) | 0.98  (0.92, 1.04) |
| Pulm. HTN | 0.98  (0.89, 1.09) | 0.98  (0.89, 1.09) | 0.99  (0.90, 1.12) | 1.01  (0.91, 1.12) | 0.98  (0.88, 1.10) |
| PVD | 0.92  (0.87, 0.98) | 0.94  (0.88, 0.99) | 0.94  (0.88, 1.00) | 0.92  (0.87, 0.98) | 0.91  (0.86, 0.98) |
| HTN | 0.99  (0.95, 1.03) | 1.00  (0.95, 1.04) | 1.00  (0.96, 1.05) | 1.01  (0.97, 1.06) | 1.01  (0.96, 1.07) |
| HTN with Complications | 0.91  (0.85, 0.97) | 0.91  (0.86, 0.97) | 0.91  (0.85, 0.97) | 0.91  (0.85, 0.97) | 0.91  (0.84, 0.97) |
| Paralysis | 2.99  (2.77, 3.23) | 2.95  (2.72, 3.19) | 2.94  (2.72, 3.19) | 2.93  (2.70, 3.18) | 2.89  (2.65, 3.16) |
| Neurological Disorders | 5.28  (5.05, 5.53) | 5.33  (5.09, 5.58) | 5.33  (5.09, 5.59) | 5.26  (5.00, 5.52) | 5.09  (4.83, 5.37) |
| COPD | 0.96  (0.91, 1.00) | 0.95  (0.91, 1.00) | 0.96  (0.91, 1.00) | 0.97  (0.93, 1.02) | 0.99  (0.94, 1.04) |
| Diabetes | 0.87  (0.82, 0.92) | 0.88  (0.83, 0.93) | 0.89  (0.84, 0.94) | 0.89  (0.84, 0.95) | 0.91  (0.86, 0.97) |
| Diabetes with Complications | 0.79  (0.73, 0.86) | 0.79  (0.83, 0.85) | 0.78  (0.72, 0.85) | 0.78  (0.72, 0.85) | 0.77  (0.70, 0.84) |
| Hypothyroidism | 1.11  (1.05, 1.16) | 1.10  (1.05, 1.16) | 1.09  (1.04, 1.15) | 1.09  (1.03, 1.15) | 1.10  (1.04, 1.16) |
| Renal Disease | 0.58  (0.53, 0.64) | 0.58  (0.53, 0.63) | 0.58  (0.53, 0.63) | 0.59  (0.53, 0.65) | 0.58  (0.53, 0.65) |
| Liver Disease | 0.93  (0.86, 1.01) | 0.93  (0.86, 1.01) | 0.94  (0.86, 1.02) | 0.92  (0.85, 1.01) | 0.95  (0.86, 1.04) |
| Peptic Ulcer Disease | 1.01  (0.80, 1.27) | 1.01  (0.80, 1.27) | 0.96  (0.76, 1.21) | 1.00  (0.79, 1.27) | 0.99  (0.77, 1.27) |
| HIV | 1.01  (0.70, 1.45) | 1.02  (0.71, 1.48) | 1.07  (0.74, 1.55) | 1.04  (0.72, 1.52) | 1.07  (0.72, 1.58) |
| Lymphoma | 1.24  (1.08, 1.42) | 1.18  (1.03, 1.36) | 1.19  (1.03, 1.38) | 1.21  (1.04, 1.40) | 1.18  (1.01, 1.38) |
| Metastatic Cancer | 0.33  (0.29, 0.38) | 0.34  (0.30, 0.39) | 0.34  (0.30, 0.39) | 0.35  (0.31, 0.41) | 0.39  (0.34, 0.45) |
| Solid Tumor | 0.86  (0.81, 0.91) | 0.86  (0.81, 0.92) | 0.88  (0.82, 0.93) | 0.89  (0.83, 0.95) | 0.89  (0.83, 0.95) |
| Rheumatic Disorders | 1.25  (1.17, 1.33) | 1.26  (1.18, 1.35) | 1.26  (1.19, 1.35) | 1.26  (1.18, 1.35) | 1.27  (1.19, 1.36) |
| Coagulopathy | 0.85  (0.78, 0.93) | 0.85  (0.78, 0.93) | 0.85  (0.78, 0.93) | 0.86  (0.78, 0.94) | 0.84  (0.76, 0.92) |
| Obesity | 0.83  (0.79, 0.89) | 0.85  (0.80, 0.90) | 0.86  (0.80. 0.91) | 0.86  (0.81, 0.92) | 0.87  (0.81, 0.93) |
| Weight Loss | 1.65  (1.55, 1.76) | 1.62  (1.52, 1.73) | 1.63  (1.52, 1.74) | 1.58  (1.48, 1.70) | 1.54  (1.43, 1.66) |
| Fluid and Electrolytes Disorders | 0.79  (0.74, 0.84) | 0.80  (0.76, 0.85) | 0.80  (0.75, 0.85) | 0.81  (0.76, 0.86) | 0.81  (0.76, 0.86) |
| Blood Loss | 0.70  (0.61, 0.79) | 0.69  (0.61, 0.79) | 0.70  (0.62, 0.80) | 0.72  (0.63, 0.82) | 0.73  (0.63, 0.83) |
| Anemia | 1.05  (1.00, 1.11) | 1.06  (1.00, 1.11) | 1.06  (1.00, 1.11) | 1.05  (1.00, 1.11) | 1.06  (1.00, 1.12) |
| Alcohol Abuse | 0.83  (0.73, 0.93) | 0.83  (0.74, 0.94) | 0.84  (0.74, 0.95) | 0.85  (0.75, 0.97) | 0.85  (0.74, 0.97) |
| Drug Abuse | 0.82  (0.71, 0.94) | 0.82  (0.71, 0.95) | 0.82  (0.71, 0.95) | 0.82  (0.71, 0.96) | 0.81  (0.69, 0.95) |
| Psychoses | 0.82  (0.77, 0.87) | 0.82  (0.76, 0.87) | 0.82  (0.77, 0.88) | 0.83  (0.78, 0.89) | 0.84  (0.78, 0.90) |
| Depression | 0.98  (0.92, 1.03) | 0.98  (0.93, 1.04) | 0.98  (0.93, 1.03) | 0.97  (0.92, 1.03) | 0.98  (0.93, 1.05) |
| *ALS Symptom History* |  |  |  |  |  |
| Never Had ALS Symptoms | Reference | Reference | Reference | Reference | Reference |
| Had 1+ ALS Symptoms | 4.49  (3.97, 5.07) | 4.20  (3.71, 4.75) | 3.98  (3.51, 4.50) | 3.56  (3.14, 4.04) | 3.08  (2.70, 3.51) |
| *Urban Residence* |  |  |  |  |  |
| Lives in Non-Metropolitan County | Reference | Reference | Reference | Reference | Reference |
| Lives in Metropolitan County | 1.42  (1.27, 1.60) | 1.42  (1.27, 1.59) | 1.42  (1.26, 1.59) | 1.41  (1.26, 1.58) | 1.40  (1.25, 1.58) |
| *Interaction Between Residency and Symptoms* |  |  |  |  |  |
| Lives in Metropolitan County **and** has ALS Symptoms | 0.83  (0.73, 0.95) | 0.83  (0.73, 0.95) | 0.83  (0.73, 0.95) | 0.84  (0.73, 0.96) | 0.85  (0.74, 0.97) |
| **Model Information** |  |  |  |  |  |
| Number of Observations | 113,827 | 112,410 | 111,062 | 107,147 | 98,850 |
| Number of Cases of ALS | 18,342 | 17,615 | 17,016 | 15,574 | 13,418 |
| C statistic | 0.795 | 0.794 | 0.793 | 0.789 | 0.778 |
| Pseudo R^2^ | 0.150 | 0.146 | 0.142 | 0.133 | 0.117 |
